# Prospective Clinical Evaluation of a Deep Learning Algorithm for Guided Point-of-Care Ultrasonography Screening of Abdominal Aortic Aneurysms

**DOI:** 10.1101/2024.02.06.24302423

**Authors:** I-Min Chiu, Tien-Yu Chen, You-Cheng Zheng, Xin-Hong Lin, Fu-Jen Cheng, David Ouyang, Chi-Yung Cheng

## Abstract

**Background:** Abdominal Aortic Aneurysm (AAA) is a critical condition that can lead to fatal consequences if not detected and treated early. Despite the high prevalence in smokers and guideline recommendation for screening, AAA often remains undetected due to availability of diagnostic ultrasound examinations. This prospective clinical trial aimed to investigate the use of a Deep Learning (DL) algorithm to guide AAA screening.

**Methods:** This prospective, comparative diagnostic study was conducted at the Kaohsiung Chang Gung Memorial Hospital. We developed and deployed an object detection-based DL algorithm providing real-time guidance for novice users performing AAA screening using point of care ultrasound. 10 registered nurses with no prior ultrasonography experience were recruited and performed at least 15 scans on patients over 65 years old to acquire abdominal aorta videos. These scans were compared with those of physicians using the same ultrasound hardware but without DL guidance.

**Results:** A total of 184 patients (median [IQR] age of 72 [67-79], and 105 (57.1%) male) completed this study. The DL-guided novices achieved adequate scan quality in 87.5% (95% CI: 82.7 - 92.3%) of patients, comparable to the 91.3% (95% CI: 87.2-95.4%) rate of physician scans (p=0.310). This performance did not vary by BMI. The DL model predicted AAA with an AUC of 0.975, showing 100% sensitivity and 94.3% specificity. The DL model predicted the maximal width of abdominal aorta with mean absolute error of 2.8mm compared to physician measurements. 3 AAA with maximal width of aorta > 3cm were found in this study cohort.

**Conclusion:** DL-guided POCUS is an effective tool for AAA screening, providing comparable performance to experienced physicians. The use of this DL system could democratize AAA screening and improve access, thereby aiding in early disease detection and treatment.

**Clinical Perspective:** *What is New:*

- Our study presents a deep learning (DL) guidance system that enables novice users to perform Abdominal Aortic Aneurysm (AAA) screening with POCUS, yielding image quality comparable to experienced physicians.
- The DL algorithm accurately identifies AAA from scans conducted by novice users, maintains consistent performance across patients with varying BMIs, and demonstrates increased scan efficiency with repeated use.

*Clinical Implications:*

- DL-guided POCUS can potentially expand AAA screening capabilities to non-specialist settings and increase throughput for screening at risk individuals.
- The implementation of our DL model for AAA screening could enhance early detection, particularly in underserved areas, but also optimize clinical workflows by decreasing diagnostic wait times and increasing ultrasound utilization efficiency.

## Introduction

An abdominal aortic aneurysm (AAA) is the gradual dilation of the abdominal aorta, and if left untreated, it may rupture, posing a high risk of fatal consequences^1^. This life-threatening condition contributes to a crude mortality rate of approximately 150,000–200,000 deaths per year worldwide^2^. More than two-thirds of patients with a ruptured abdominal aortic aneurysm present without a prior diagnosis of AAA^3^. Early detection and treatment can considerably decrease AAA-related mortality, especially in the elderly ^4^. Previous guidelines recommend ultrasonography for general AAA screening in at-risk populations, specifically men over 65, with smoking history^5,6^.

Ultrasonography, the most common imaging screening modality for AAA, has proven to be effective and less costly compared to standard computed tomography (CT) scans.^7^. Nonetheless, ultrasound is generally performed by highly trained sonographers and interpretation carried out by board-certified physicians. This challenge is further compounded by the restricted accessibility of sophisticated ultrasound equipment. Both of these factors contribute to ultrasound examinations having the longest waiting times compared to other imaging modalities and reduce the cost-effectiveness for general screening for AAA^8–10^.

In the past decade, Point-of-Care Ultrasound (POCUS) has seen extensive use across various hospital settings, including outpatient clinics, emergency departments, wards, and operating rooms^11,12^. It has played a crucial role in medically underserved areas, from rural regions in developed countries to low-income nations, and is even used in manned space flights^13^. Portable ultrasound machines have facilitated the acquisition of high-quality images suitable for diagnostic purposes^14^. However, the limited familiarity among hospital staff with ultrasound its current utilization^15^. Therefore, integrating AI guidance into ultrasound could benefit novice operators. This has the potential to enhance diagnostic capabilities, especially in remote areas where experienced sonographers may not be readily available.

AI revolutionizes healthcare by rapidly interpreting images, detecting abnormalities, segmenting organs and lesions, and facilitating early disease identification. Prior study suggests that AI empowers individuals with no previous experience in ultrasonography to perform diagnostic transthoracic echocardiographic studies, encompassing the evaluation of left and right ventricular function, as well as the identification of pericardial effusion^21^. In addition, AI can guide novices to capture satisfactory diagnostic images of the Morison pouch during Focused Assessment With Sonography in Trauma (FAST) exam^22^. The objective of this study is to develop and validate a Deep Learning (DL) model that guides abdominal aorta scanning and to investigate its potential in assisting novices to obtain qualified scans. This approach could aid in the screening of potential AAA patients by advancing the detection of symptomatic aneurysms, screening asymptomatic AAAs in at-risk groups, and monitoring aneurysm growth until treatment is necessary.

## Method

### Ethical Approval

The study was approved by the institutional review board at the Kaohsiung Chang Gung Memorial Hospital (IRB number: 202102311B0). Written consent was obtained from each participant.

### Development and Function of AI-Guided Image Acquisition Software

The DL-guided image acquisition algorithm is detailed in the Supplemental Methods. It offers real-time, continuous guidance during scanning to assist users in obtaining videos for AAA screening. The software, which emulates physician expertise, utilizes a You Only Look Once (YOLO) architecture, known for its real-time object detection capabilities. It is specifically tailored to analyze ultrasonographic images, focusing on identifying anatomical structures including the abdominal aorta, spine, and inferior vena cava (Supplemental Figure S1).

The algorithm operates solely based on ultrasonographic images, eliminating the need for external trackers, fiducial markers, or additional sensory inputs. This streamlines the diagnostic workflow and enhances screening efficiency. In this research, we installed the software on a commercially available POCUS system (Telemed MicrUs EXT-1H). After integration, the software monitors and processes the ultrasound display continuously through its application programming interface (Supplemental Figure S2). It detects and provides the anatomical location of the abdominal aorta on the ultrasound display, thereby guiding users to improve image acquisition during scanning.

The DL algorithms were trained using 2,101 POCUS images focusing on AAA screening from the studied hospital and externally validated in 492 images from a local hospital. These images were retrospectively collected from routine scans conducted in the Emergency Departments (EDs) of both hospitals. The performance of the algorithm achieved an average precision of 0.973 in internal validation and 0.843 in external validation. Supplemental Methods depicts the DL model training data set, expert labeling for image quality, algorithm optimization, and integration of DL system to POCUS hardware in the study.

### Study Design

Patients at least 65 years old visiting the outpatient clinic of the Cardiology department at the studied hospital were recruited between June and August 2023. Individuals were excluded if they were unable to lie flat or were unable or unwilling to provide informed consent.

10 registered nurses without prior experience performing or interpreting ultrasonography were recruited for the trial from hospital personnel. Each nurse underwent a 15-minute tutorial to become familiarized with the POCUS machine and DL guidance. After that, they were asked to perform 15-20 scans to acquire a 10-second standard abdominal aorta tracing video under DL guidance. For control, a duplicate scan was obtained by a physician using the same POCUS machine on the same day but without AI guidance. The physician also labeled the maximal width of the aorta for the control scan. The nurse scans were conducted independently, solely with DL guidance, and always preceded the control scans. Following each scan, the Telemed POCUS machine stored two ultrasonography videos at 20 frames per second, which the DL system then processed to predict the maximal width of the abdominal aorta. Figure 1 illustrates the study design.

**Figure 1.**
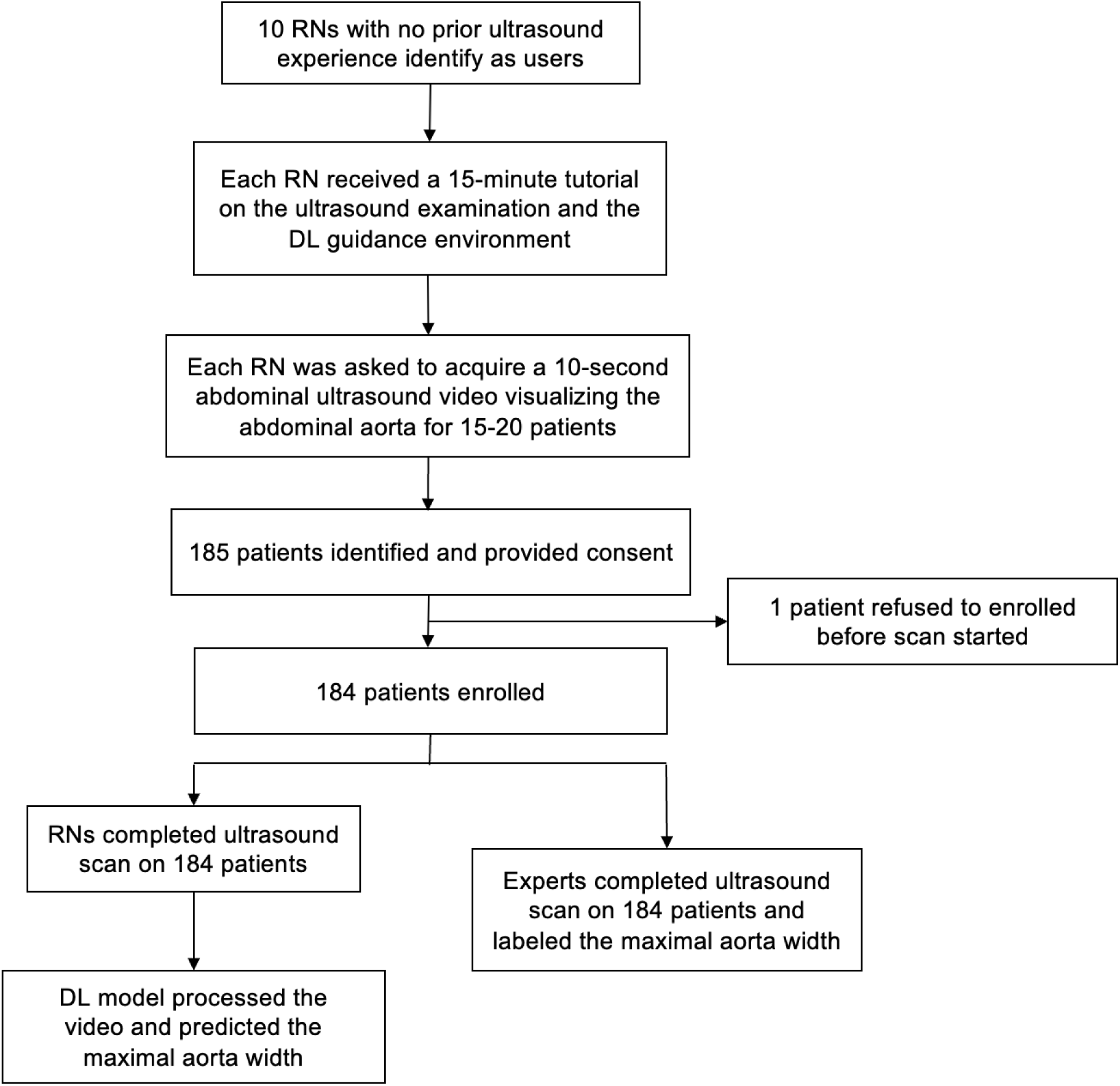
Study design.

Upon completion of all study and control scans, a panel of 3 expert physicians (Y.C.C, X.H.L., and F.J.C.) independently and blinded to whether a nurse or a physician performed the study, assessed whether each scan was of diagnostic quality, served as the primary endpoint. All expert readers were certified board physicians in Cardiology or Emergency Medicine. The time to complete the study, defined as the interval from placing the probe on the patient’s abdomen to completing the scan, was recorded. The maximal aortic width predicted by the DL model was compared with expert measurements for the secondary endpoint.

### Statistical Analysis

The study sought to evaluate the performance of nurses conducting AAA screening under DL guidance, with continuous variables reported as medians and interquartile ranges (IQR), and categorical variables as numbers and percentages. The proportion of qualified studies as judged by the expert panel was compared between DL guidance and physician scans for the primary endpoints. The maximal abdominal aortic width measurement and time to complete the study were evaluated as secondary endpoints. For both primary and secondary parameters, the proportion judged clinically evaluable is reported with 95% confidence intervals (CIs).

## Result

During the study period, 185 patients were included. 1 patient refused to be enrolled before the scan started, while 184 patients completed both the nurse and physician examinations (Figure 1). Their median (IQR) age was 72 (67-79), 57.1% were male, and the median (IQR) Body Mass Index (BMI) was 25.1 (23.3-28.1). 131 (71.2%) of them had Hypertension, 64 (34.8%) had Diabetes Mellitus, 83 (45.1%) had heart disease, and 41 (22.3%) had a smoking history. AAA was diagnosed by physicians in 3 patients, representing 1.6% of the study cohort. Table 1 shows the demographics of the included patients.

**Table 1.**
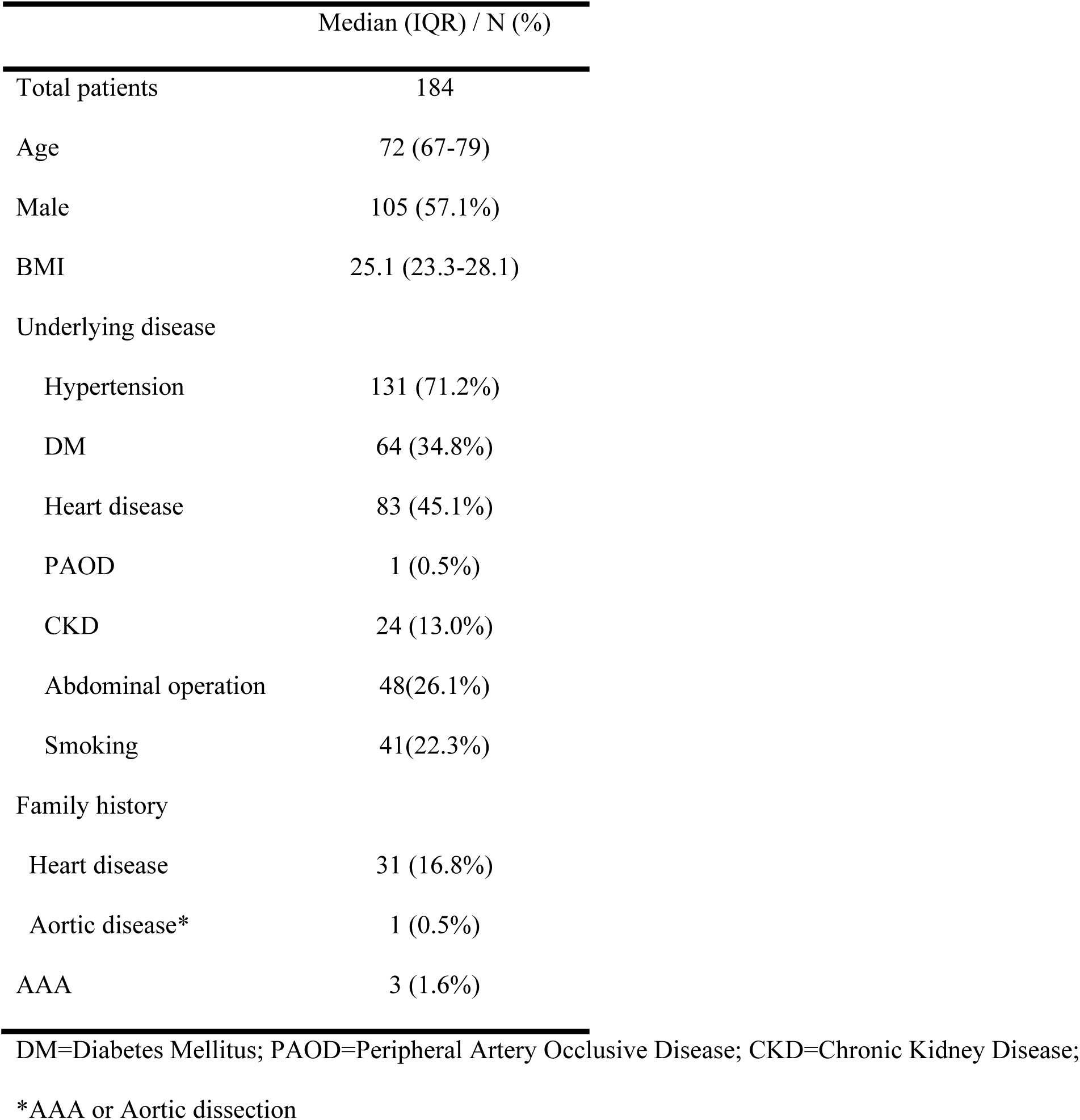
Demographic of included patients.

|  | Median (IQR) / N (%) |
| --- | --- |
| Total patients | 184 |
| Age | 72 (67-79) |
| Male | 105 (57.1%) |
| BMI | 25.1 (23.3-28.1) |
| Underlying disease |  |
| Hypertension | 131 (71.2%) |
| DM | 64 (34.8%) |
| Heart disease | 83 (45.1%) |
| PAOD | 1 (0.5%) |
| CKD | 24 (13.0%) |
| Abdominal operation | 48(26.1%) |
| Smoking | 41(22.3%) |
| Family history |  |
| Heart disease | 31 (16.8%) |
| Aortic disease* | 1 (0.5%) |
| AAA | 3 (1.6%) |

DM=Diabetes Mellitus; PAOD=Peripheral Artery Occlusive Disease; CKD=Chronic Kidney Disease;
\*AAA or Aortic dissection

Regarding primary outcome, no significant difference was found in the rate of qualified videos between DL-guided scans 87.5% (95% CI: 82.7 - 92.3%), and physician-performed scans 91.3% (95% CI: 87.2-95.4%), with p value of 0.310. Furthermore, the qualified rate for DL guidance remained consistent across patients with varying BMI levels: 87.6% (95% CI: 80.9 - 94.1%) in patients with BMI > 25 and 87.4% (95% CI: 80.4 - 94.3%) in patients with BMI < 25. After 5 rounds of examination, the proficiency of scans slightly increased, reaching an 88.1% (95% CI: 82.3 - 93.9%) qualification rate, as detailed in Table 2.

**Table 2.** Comparison of Nurse-Acquired and Physician-Acquired Studies for Primary and Secondary Clinical Parameters.

|  | Physician | DL guidance | p-value |
| --- | --- | --- | --- |
| Qualified Video | No. (%) [95% CI] |  |  |
| Total | 168 (91.3) [87.2 - 95.4] | 161 (87.5) [82.7 - 92.3] | 0.310 |
| BMI>25 (N=97) | 91 (93.8) [87.6 - 99.8] | 85 (87.6) [80.9 - 94.1] | 0.137 |
| BMI<25 (N=87) | 77 (88.5) [81.5 - 95.4] | 76 (87.4) [80.4 - 94.3] | 0.816 |
| Before 5 scans (N=50) | 45 (90.0%) [81.7-98.3] | 43 (86.0%) [76.4-95.6] | 0.613 |
| After 5 scans (N=134) | 123 (91.8) [86.5 – 97.1] | 118 (88.1) [82.3 - 93.9] | 0.659 |
| Time to Complete Study | Median (IQR), Seconds |  |  |
| total | 20 (16-33) | 37 (21-60) | <0.001 |
| BMI>25 (N=97) | 21 (16-35) | 42 (27-67) |  |
| BMI<25 (N=87) | 18 (15-28) | 30 (18-54) |  |
| Before 5 scans (N=50) | 20 (16-35) | 53 (38-82) |  |
| After 5 scans (N=134) | 20 (15-30) | 30.5 (18-55) |  |

The time to complete the study was longer with DL guidance, averaging 37 seconds (IQR 21-60), compared to 20 seconds (IQR 16-33) for physician-led scans (p<0.001). For nurses using DL guidance, the completion time was longer in patients with a BMI over 25, taking 42 seconds (IQR 27-67), as opposed to 30 seconds (IQR 18-54) for those with a BMI under 25. Physicians’ scan showed similar pattern between different level of BMI. With increased use of the DL system, nurses’ scan times decreased; after five scans, the average time was reduced from 53 seconds (IQR 38-82) to 30.5 seconds (IQR 18-55) as reported in Table 2.

Of the 161 scans under DL guidance classified as diagnostic quality, the predicted maximal aortic widths showed a mean absolute error of 2.8mm compared with physician measurements, as depicted in Figure 2. Of these scans, 159 (98.8%) had a discrepancy of less than 1cm when compared to physician labeling. Three patients (1.6%) were diagnosed with AAA in this study based on the physician’s ultrasound findings. The DL model can predict AAA with an AUC of 0.975 (95% CI: 0.943-1). While using 2.5cm as the cut-off threshold, the DL model has 100% sensitivity in detecting AAA in these patients, along with 94.3% specificity, 33.3% positive predictive value, and 100% negative predictive value (Table 3).

**Figure 2.**
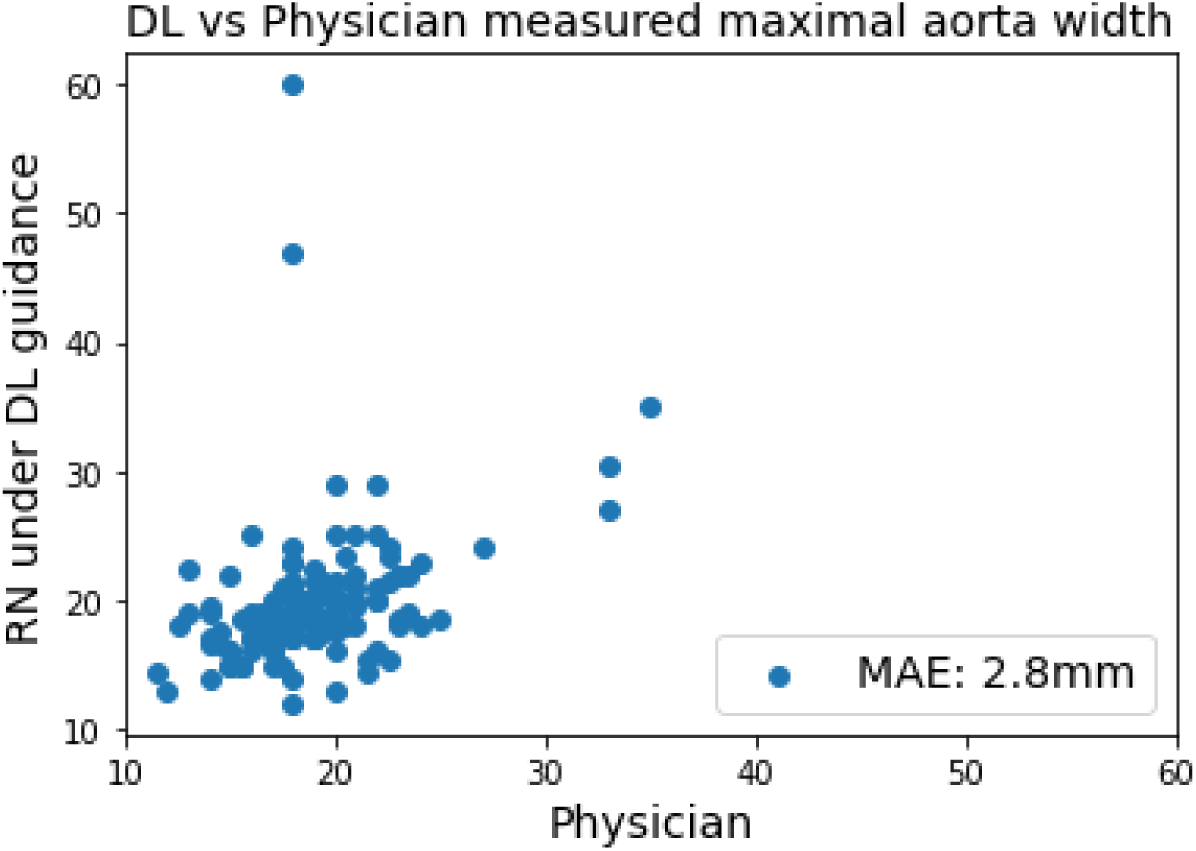
Scatter Plot of Maximal Aortic Width Measurements by Deep Learning Predictions and Physician Labels.

**Table 3.**
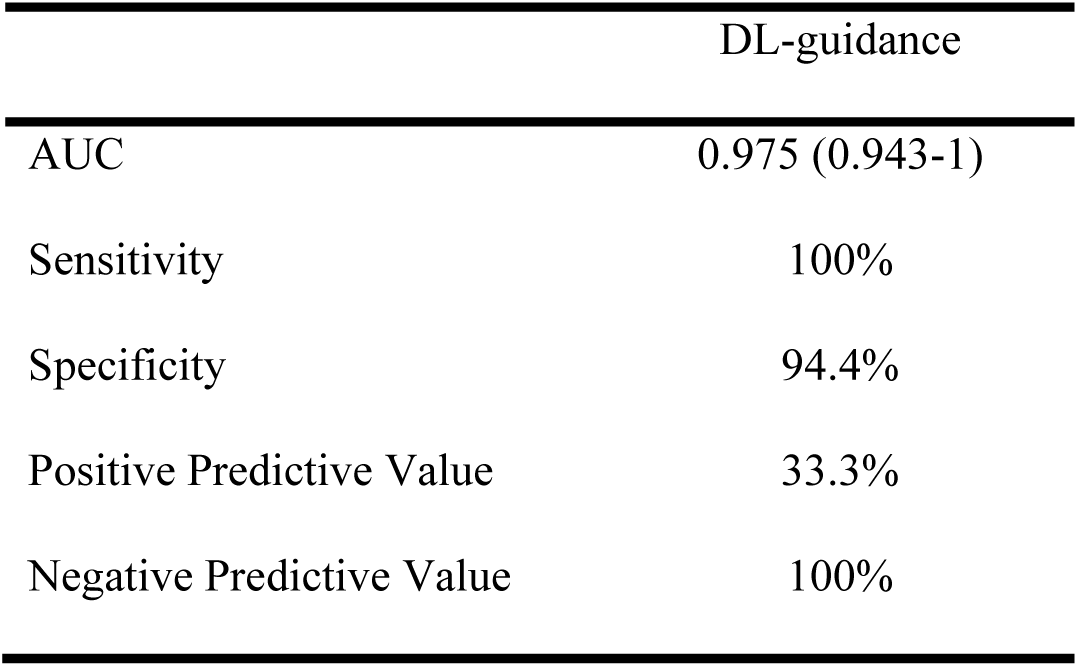
Diagnostic performance for predicting AAA by DL algorithm.

## Discussion

In this study, we developed and validated a DL algorithm for guiding novice users performing AAA screening with POCUS, which notably demonstrated parity with experienced physicians in producing diagnostic-quality videos. This finding suggests DL guidance can compensate for a lack of extensive sonography experience, potentially broadening AAA screening accessibility. Moreover, the DL model exhibited robust performance across a range of patient body mass indexes and showed a notable learning curve, with improved scan times after repeated use. The precision of our DL model in predicting the presence of AAA was quantitatively reflected by an AUC of 0.973 in our sample population. This high level of accuracy not only demonstrates the DL model’s capability to distinguish between normal and pathological findings but also suggests its potential as a reliable tool for early detection. These results underscore the practicality and efficiency of implementing AI in clinical ultrasound practice, that may help to reduce waiting time for ultrasound examination especially in resource-limited settings where access to skilled sonographers is challenging.

The utility of POCUS is well recognized for its convenience and the immediacy with which it delivers diagnostic imaging at the patient’s bedside. However, its effective use is traditionally limited by the operator’s expertise, with the quality of the interpretation being heavily reliant on the sonographer’s experience. Current protocols require at least a 3-month training program for technicians to become familiar with examining the abdominal aorta using ultrasound^23^. This presents a significant challenge in resource-limited settings where access to highly trained professionals is scarce^24,25^, which led to ongoing debate about the cost-effectiveness and relevance of AAA screening. Given its low prevalence, with historical data shows the prevalence rate of AAA 1.3% to 4.9 in selected risk population, the balance between the costs and benefits of widespread screening is called into question^26,27^. This is precisely where the integration of DL guidance in AAA screening could play a transformative role. By potentially reducing the time and expertise required for accurate screening, DL guidance can lower operational costs and improve the efficiency of screening programs. Moreover, the enhanced accuracy of DL-guided screening might lead to more effective identification of AAA cases even in a landscape of lower prevalence, ensuring that resources are optimally utilized.

Advances in cardiovascular ultrasound interpretation using AI have been significant in recent years, with numerous studies demonstrating automated quantification of cardiac structures and function, and AI-driven disease identification showing less variability than semi-automated or manual analyses^28–31^. The convergence of AI-guided acquisition with automated interpretation could expand ultrasound’s reach, improving the recognition of pathology. Our study’s DL algorithm addresses this by providing real-time guidance to novice users, effectively bridging the gap between the ease of use of POCUS and the expertise required for accurate diagnosis. By supplementing the user’s limited experience with sophisticated AI, we facilitate a higher standard of care that could potentially revolutionize the screening process for conditions like AAA, where early detection is crucial.

Several potential explanations for our findings emerge upon examination of the DL algorithm’s performance. The ability of the algorithm to offer real-time, continuous guidance likely played a professional guiding role in enabling novice operators to achieve a high rate of qualified videos. This real-time feedback is particularly useful during scanning, as it assists users in adjusting the ultrasound probe to optimize the visualization of the aorta. Such immediate guidance can improve image quality, a benefit observed in previous studies where AI support enhanced the outcomes of sonography procedures, such as in the Echocardiography^21^ or Focused Assessment With Sonography in Trauma exams^22^. The similarity between these findings and our own suggests a shared advantage of AI assistance across different ultrasound applications. The learning curve evidenced by the reduction in scan times with repeated use indicates that users are not only becoming more adept at operating the AI system but are also gaining a better understanding of the aortic structure. This suggests a synergistic enhancement in the operator’s ability to acquire diagnostic images, pointing to a productive interaction between human users and the AI tool over time.

The architecture of our DL model, which delineates the abdominal aorta with a bounding box in each frame of the ultrasound video, serves two critical functions. First, it offers real-time feedback during the scanning process, guiding users in adjusting their probe to achieve optimal imaging. Second, it enables precise diagnostic measurements post-scan. By accurately capturing the anatomy of the aorta, our algorithm processes each frame to determine the aortic width, subsequently calculating the maximal width from the entire video. The minimal average error of 2.8mm between the expert measurements and those obtained via DL guidance, along with the high AUC for detecting an AAA, attest to the DL model’s effective training. For further evaluation, a threshold of 2.5cm led to expert review for 9 (4.9%) of the 184 scans guided by DL. Within this group, three AAA cases were accurately identified, resulting in a sensitivity of 1.0 and a specificity of 0.94 in our cohort (Table 3). Notably, there were two scans where the discrepancy in maximal width measurements between expert interpretation and DL guidance exceeded 1cm (Figure 2). Manual review of these outliers showed that the DL algorithm had incorrectly identified a 6.0 cm liver cyst and a 4.8 cm fluid-filled small bowel loop. These cases of misclassification demonstrate that when videos with predictive diameters suspicious for AAA are scrutinized, physicians can readily discern true positives from false positives.

## Limitations

There are a few limitations of the study. First, the study was conducted in a controlled clinical setting in the outpatient clinic, which allowed access to a large patient population but may not fully replicate POCUS use conditions in remote or underserved areas. Second, while the sample size was sufficient for the primary endpoints, the relatively small number of patients and nurses limits the assessment of generalizability; larger studies could yield more robust data. Additionally, there was no control group for the nurse scanners. The comparison in the study was against physician’s acquisitions, but an additional control group of novices untrained with the algorithm was not used. Furthermore, the study location was one tertiary academic hospital.

Further validation across a variety of settings would help strengthen the results of the study. The DL model’s performance in a real-world screening scenario may differ from our controlled environment. Notwithstanding these limitations, the strengths of the study included a rigorous methodology, integration of the DL to the use of a commercially available POCUS system.

## Conclusion

In conclusion, our study indicates that a DL-guided POCUS can serve as an effective tool for AAA screening, achieving diagnostic accuracy that is on par with experienced physicians. This innovative approach has the potential to democratize AAA screening, enhancing accessibility and cost-effectiveness. By harnessing the capabilities of AI, we can streamline the screening process, reduce the need for extensive sonographic training, and potentially improve patient outcomes through early detection.

## Data Availability

Not available

## Supplemental Methods

### Data Curation

For developing the DL model, we collected ultrasound images from the ultrasound machines, include Sonosite Edge II and Hitachi Noblus, in the ED of Kaohsiung Chang Gung Hospital from Jan 2019 to Dec 2021. Ultrasound images focusing on abdominal area were collected. Images that were not related to abdominal aorta examination were excluded and eventually a dataset comprising 2,101 labeled ultrasound images was used. We also collected 492 ultrasound images from a regional hospital for external validation.

Each ultrasound image was cropped into 600*400 in size and get rid of the information that may reveal personal identification. Two medical experts on point of care ultrasound then manually labeled the selected anatomical structures - the aorta, inferior vena cava (IVC), and spine (Figure S1) with polygon mask. We adopted 6 commonly used labeling software, Labelme, for this study and save the labeling file under COCO dataset format. This large, annotated dataset served as the foundation for training the AI models, enabling them to recognize and correctly identify these structures in ultrasound images.

### DL Model Development and Validation

Two DL models were developed. The first was an object-detection-based AI model aim to provide real-time feedback and the second was trained to automatically identify and outline the aorta, inferior vena cava, and spine within each frame of the ultrasound video. These models were trained and initially validated using the annotated dataset, reserving a portion for later testing. The first architecture employed in our study is YOLOv5 instance segmentation. The input is an ultrasound image with a resolution of 600 x 600, and the inference output includes bounding boxes and pixel area identification for each category (abdominal aorta, spine, and inferior vena cava). As the original ultrasound images vary in size, we opted for the ‘letterbox’ preprocessing method to standardize them to the model’s required dimensions. Letterboxing scales the image while maintaining the original aspect ratio; any remaining space after scaling is filled with the background, mimicking the effect of placing a picture into an envelope, hence the term ‘letterbox’. YOLOv5, in terms of its architecture, originates from YOLOv4’s CSPDarknet53, and incorporates several improvements to enhance speed and inference outcomes. For instance, it replaces the Spatial Pyramid Pooling (SPP) with SPPF to boost computational speed and utilizes techniques such as Copy-Paste for data augmentation. The result of validation was shown in Table S1.

### Integrated DL model with POCUS

The POCUS equipment used in this study is the ArtUs-EXT-1H from Telemed, a FDA-certified platform for capturing raw ultrasound signals. It can be used in conjunction with a portable tablet computer. The tablet runs on a Windows 11 environment and uses an Intel CPU (detailed specifications are listed below). To accelerate the inference speed of the deployed model, the trained YOLOv5 model was converted from the PyTorch (.pt) format to the OpenVINO IR (FP16) format and utilized via OpenVINO Runtime. We used Python’s built-in ctypes library to load the dll (dynamic link library) provided by Telemed, allowing real-time ultrasound images to be captured within the Python program. The program also allows for model inference and uses OpenCV to visually represent the identified Aorta (Figure S2). Additionally, to accommodate for the need to adjust parameters such as TGC (time gain compensation) during scanning, a control panel interface is displayed using Tkinter for the operator to adjust as necessary.

**Table S1.** DL model performance in internal and external validation.

|  | Box Precision | Box Recall | Box mAP@0.5 |
| --- | --- | --- | --- |
| Internal Validation | 0.921 | 0.987 | 0.973 |
| External Validation | 0.627 | 0.943 | 0.843 |

**Figure S1.**
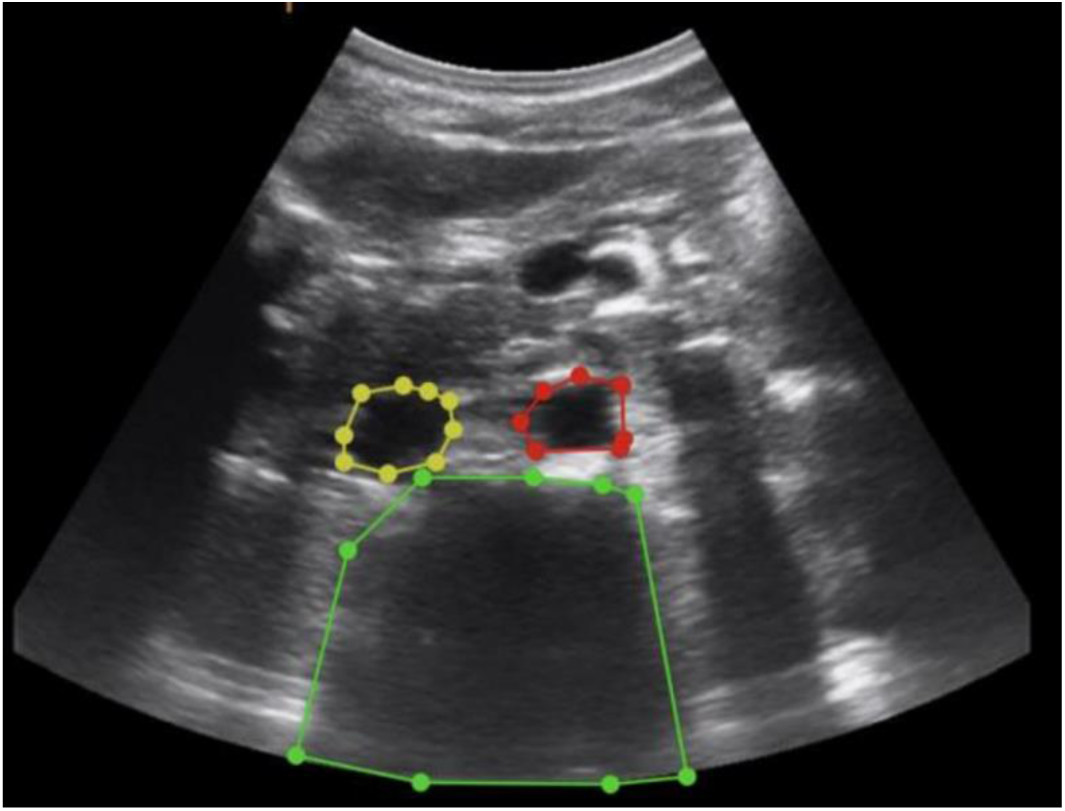
An example of label over abdominal aorta, spine, and inferior vena cava.

**Figure S2.**
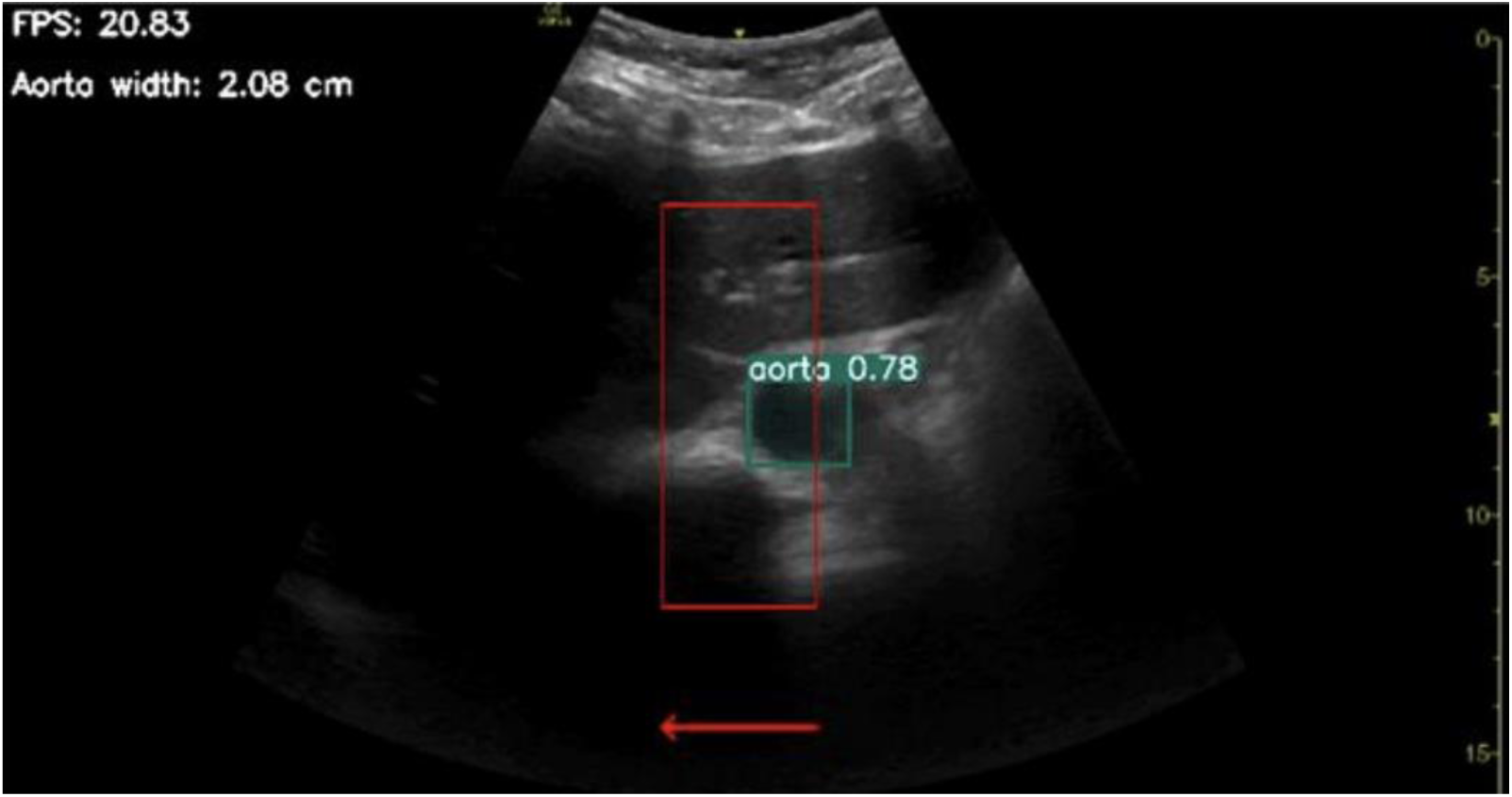
user interface of the DL guidance for image acquisition on POCUS.

